# SYSTEMATIC REVIEW EXPLORING THE IMPACT OF SOCIO-CULTURAL FACTORS ON PAIN MANAGEMENT APPROACHES IN SUB- SAHARAN AFRICA

**DOI:** 10.1101/2022.04.11.22273701

**Authors:** Henrietta Obaabeng Dompreh, Mary Lynch, Mary Longworth

## Abstract

**Aim:** The experience and expression of pain are influenced by numerous factors of which culture and the society plays a major role especially in SSA. However, few studies have focused on the impact of cultural influences on pain assessment and management in SSA. This systemic review examines pain prevalence and its intensity/severity, the socio-cultural factors that affect pain management and the extent to which socio-cultural practices influence pain assessment and management in SSA.

**Methods:** Applying the Preferred Reporting Items for Systemic Reviews and Meta-Analyses (PRISMA) guidelines, a systematic literature search was conducted. Seven electronic databases were searched, and a strict inclusion and exclusion criteria applied to the retrieve articles along with a robust filtering to identify eligible peer reviewed literature. The review process concluded with 24 eligible articles and following the Grades of Recommendation, Assessment, Development and Evaluation (GRADE) approach was applied to assess the quality of the included literature and thematic narrative analysis was conducted.

**Results:** The analysis findings identified that there are sociocultural barriers to effective pain management from the perspective of different subcultures in SSA. The evidence suggests that religious/spiritual and inherited beliefs, along with limited knowledge and health literacy influence the experience and pain management approaches applied in SSA. In addition, results indicate that, resource constraints and cultural and societal norms impact on access and use of pain management among the population in SSA.

**Conclusion:** Healthcare professionals should be aware of how the society, cultural and beliefs of their patients influence their expression of pain and subsequent pain management. Under-treatment or over-treatment might occur if health workers are unaware or do not consider the cultural norms associated with pain and pain expression, due to the subjective and individual nature of pain.

## Introduction

The disease burden of acute and chronic conditions and the risk for suffering weighs heavily on Sub Saharan African (SSA), due to the high global disease burden and pain disproportionately prevalent in sub-Saharan Africa (Young et al 2018). This has led to the concentration of efforts in SSA and developing countries to primarily focus on disease eradication, management of emerging conditions and decreasing mortality and morbidity rate. However, this health focus does not rule out the accompanied level of pain/suffering associated with disease burden, along with management of health conditions in the eradication process. The prevalence and severity of pain is not only due to disease burden, but also, results from the limited capacity to effectively diagnose pain coupled with unavailability of efficacious treatment options (Miftah *et al* 2017). Nevertheless, because the focus in SSA is on disease eradication, there has been less focus on the accompanied level of pain and requires effective pain management approaches (Young et al., 2018). Pain is a major symptom of disease and a characteristic of many medical conditions, therefore, a contributing factor for individuals seeking medical intervention.

Pain at present, has no common understanding and universally accepted definition for nor description within the health sector, despite the increasing research on pain concepts (Burton *et al*., 2017). Pain can be described as any undesirable experience associated with an actual or potential physiological changes, and not just a physiological response to tissue damage (Kumar & Elavarasi, 2016). Perceived pain is an individual’s personal experience which can be influenced by numerous factors (Callister, 2003). These causes can include the behavioral and emotional responses expected and accepted by one’s society which may influence an individual’s perception of pain (Calvillo & Flaskerud 1993). Societal cultural considerations can impact on sensitivity to pain which can make pain difficult to assess and measure (Peacock & Patel 2008). Evidence indicates that for pain to be effectively assessed and managed, requires an individual to precisely communicate the nature of pain, where pain is felt along with the severity experienced (Herr *et al*., 2011). However, factors contributing to effective pain assessment and management can be affected by the perceived beliefs and societal influence in which the individual may reside. Therefore, in aspiring to take a holistic patient centered approach in the affective management of pain, cultural and societal factors must be considered during assessment (Lasch 2000). People’s exposure to culture, society, ethnicity/tradition, largely affects the individual’s behaviour as these unique factors impact on beliefs, knowledge, practices, values, and everyday life activities (Callister 2003). Despite culture being known as a major influence on pain perception, assessment, and management Nortjé and Albertyn (2015), there is limited evidence available to date on the effect socio-culture has on pain assessment and management.

Failing to recognise the effect of socio-culture on pain management can lead to issues such as cultural conflict, miscommunication, misdiagnosis, inappropriate and ineffective patient care. Evidence to date on pain assessment and management linked with socio-cultural factors are from developed countries (Finnström & Söderhamn 2006) and little is known on the specific socio-cultural factors that impact on pain management approaches in developing counties in SSA.

The aim of this systematic review is to examine the evidence of the influence of socio-cultural factors on pain management approaches in SSA. This systemic review will examine the literature on pain prevalence, experiences of intensity/severity of pain, the socio-cultural factors that affect pain management and the extent to which socio-cultural practices impacts on pain assessment and management in SSA.

## Methods

This systematic review followed the Preferred Reporting Items for Systematic Reviews and Meta-Analyses (PRISMA) guidelines (Moher et al (2009). The inclusion criteria for eligible literature were peer reviewed articles focused on sociocultural factors and it influence on pain management approaches in SSA, that were published between 2001 and 2021 in full text articles available in English. The exclusion criteria were articles that did not focus on sociocultural factors and its influence on pain management approaches in SSA.

### Search strategy and selection criteria

The literature search strategy was conducted using the databases; CINAHL, Global Health (CABI), MEDLINE, Web of Science, PsycINFO, Cochrane Library and PubMed Central Open Access. The search strategy included manual screening of the indexes of relevant journals and reference lists of relevant primary studies. Published studies were also searched and their keywords used for the search terms. These search terms were screened independently and agreed by the researcher along with consultation with the subject Librarian and two additional researchers. In terms of the grey literature, authors who have carried out studies were contacted for ongoing studies that sought to either throw more light on previous studies or new study using different approach and methodology.

A MESH (Medical Subject Heading) term as well as keywords developed were used in searching the literature. To ensure that the correct articles were identified, search terms were divided into 3 groups: population, intervention, and outcomes. Search terms were linked with “or” Boolean operators and with “and” Boolean operators between groups in other to bring studies that contains at least one of the search terms for the review. This enabled the researcher to combine different search terms to retrieve as many as possible studies that may contain the search term relevant for the study objectives.

### Screening and Data extraction

Articles were independently screened by titles and abstracts, followed by evaluation of full-text articles by two researchers (HOD) and (MKL) independently and screening process required agreement by both reviewers on included articles meeting the inclusion and exclusion criteria. If there were disagreements these were resolved by the third reviewer (ML) and a consensus agreed. A data extraction form in Microsoft excel spreadsheet was used to assess the potential papers strategically and logically. Information sought from each article included author, title of article, journal, publication year, study population, and sample size.

### Quality appraisal

To assess the quality of the selected studies the Grading of Recommendations Assessment, Development and Evaluation working group methodology (GRADE) was used. GRADE is an assessment tool for rating the reliability of evidence strength of recommendations in health care (Guyatt et al., 2011). It consists of five categories of reasons for rating quality of evidence: risk of bias, publication bias, imprecision, inconsistencies, and indirectness. Depending on the score obtained after the GRADE scoring, the studies were ranked based as high, medium, or low by all the reviewers.

### Study registration

This study has been registered with PROSPERO with registration number CRD42020176518 and as well followed the PROSPERO guideline for conducting searches and extracting data.

## Results

### Study selection process

After a vigorous search of databases, 1892 studies were retrieved. This has been presented in Figure 1. 72 duplicates were removed. From the 1820 remaining articles, 1743 were excluded after screening the titles and abstracts. 47 articles were further excluded as they were not conducted in Sub-Saharan Africa. Further screening recognized 6 extra articles which were not primary studies. Consequently, 24 articles were submitted to complete content reading as well as critically appraised of the methodological quality to be included in this review.

**FIGURE 1.**
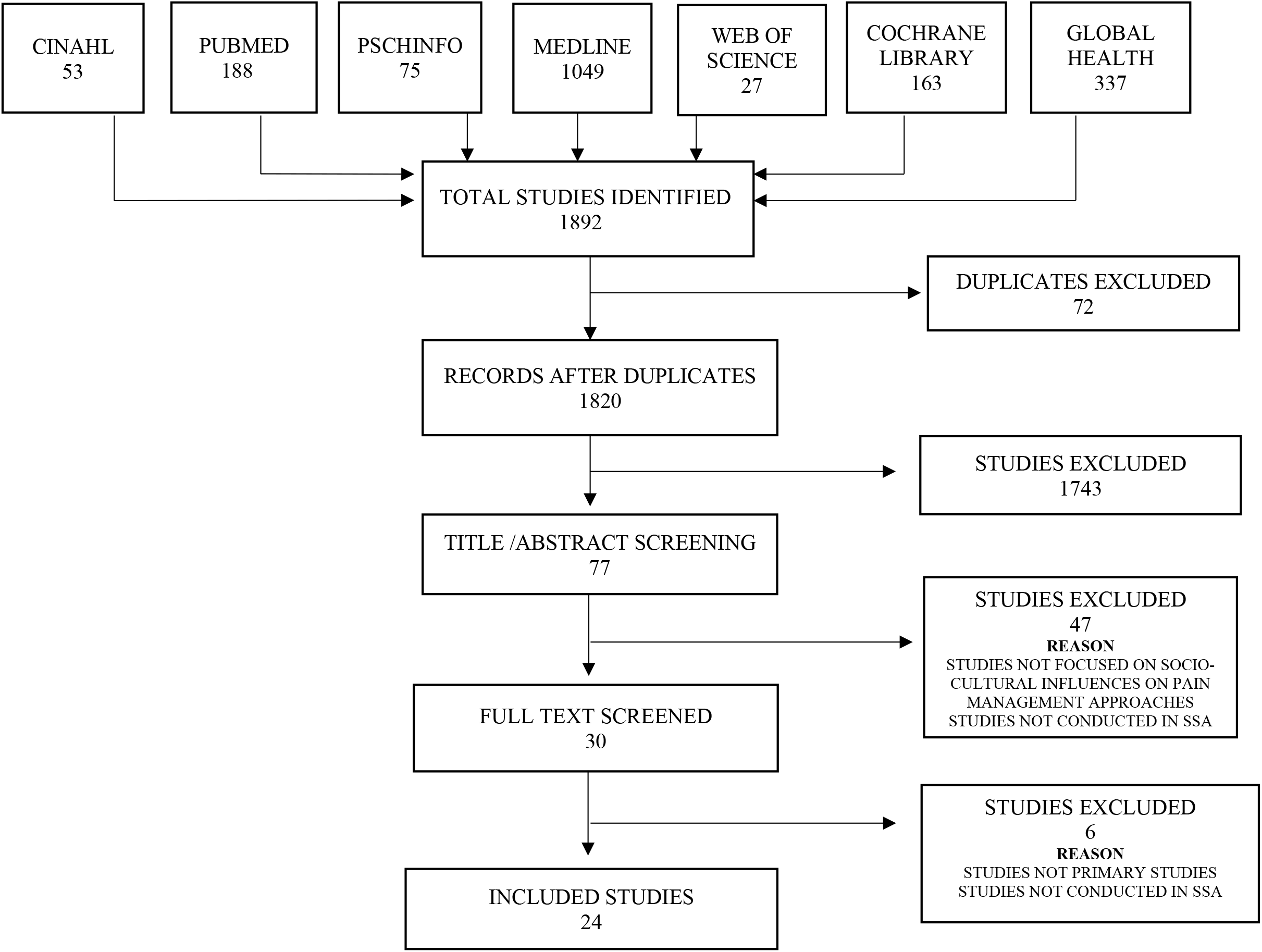
Preferred Reporting Items for Systematic Reviews and Meta-Analyses (PRISMA) flow diagram on the impact of socio-cultural factors on pain management approaches in SSA systematic review

### Characteristics and Quality assessment of selected studies

Depending on the score obtained following the GRADE scoring, the various studies were ranked based on the analysis conducted as high, medium, low, or very low by all three reviewers. Five studies were considered of high quality, 16 studies were considered of moderate quality and three studies considered as low-quality. The methodological quality of all studies was therefore appropriate and gave adequate information in terms of the findings. The characteristics and quality assessment of the included studies are outlined in Table 2.

**Table 1.** Keywords used in database searches

| PICO Framework | Keywords |
| --- | --- |
| **Population** | Boy, Youth, Juvenile, Sub-Saharan Africa, Africa, South of Sahara, Land of The Sahara, Angola, Benin, Botswana, Burkina Faso, Burundi, Cabo Verde, Cameroon, Central African Republic, Chad, Comoros, Congo, Dem. Rep., Congo, Rep., Cote D'ivoire, Equatorial Guinea, Eritrea, Eswatini, Ethiopia, Gabon, The Gambia, Ghana, Guinea, Guinea-Bissau, Kenya, Lesotho, Liberia, Madagascar, Malawi, Mali, Mauritania, Mauritius, Mozambique, Namibia, Niger, Nigeria, Rwanda, Sao Tomeand Principe, Senegal, Seychelles, Sierra, Leone, Somalia, South Africa, South Sudan, Sudan, Tanzania, Togo, Uganda, Zambia, Zimbabwe |
| **Intervention** | Socio-culture, Cultur*, Belief*, Perception, Tradition*, Art, Ideology, Religion, Society, Societal, Experience, Ethnicity, Civilization, Practice*, Custom*, Habit*, Heritage, Values, Community, Subcultural |
| **Comparator** | Not Applicable |
| **Outcome** | Pain, Discomfort, Ache, Suffering, Pain Management, Pain Relief, Pain Control, Pain responses, Analgesia*, Pain killing, Numbing, Deadening, Pain Easing, Soothing, Palliative, Assuage, Settling, Sedative, Tranquilizing, Comforting, Calmative |

**Table 2.**
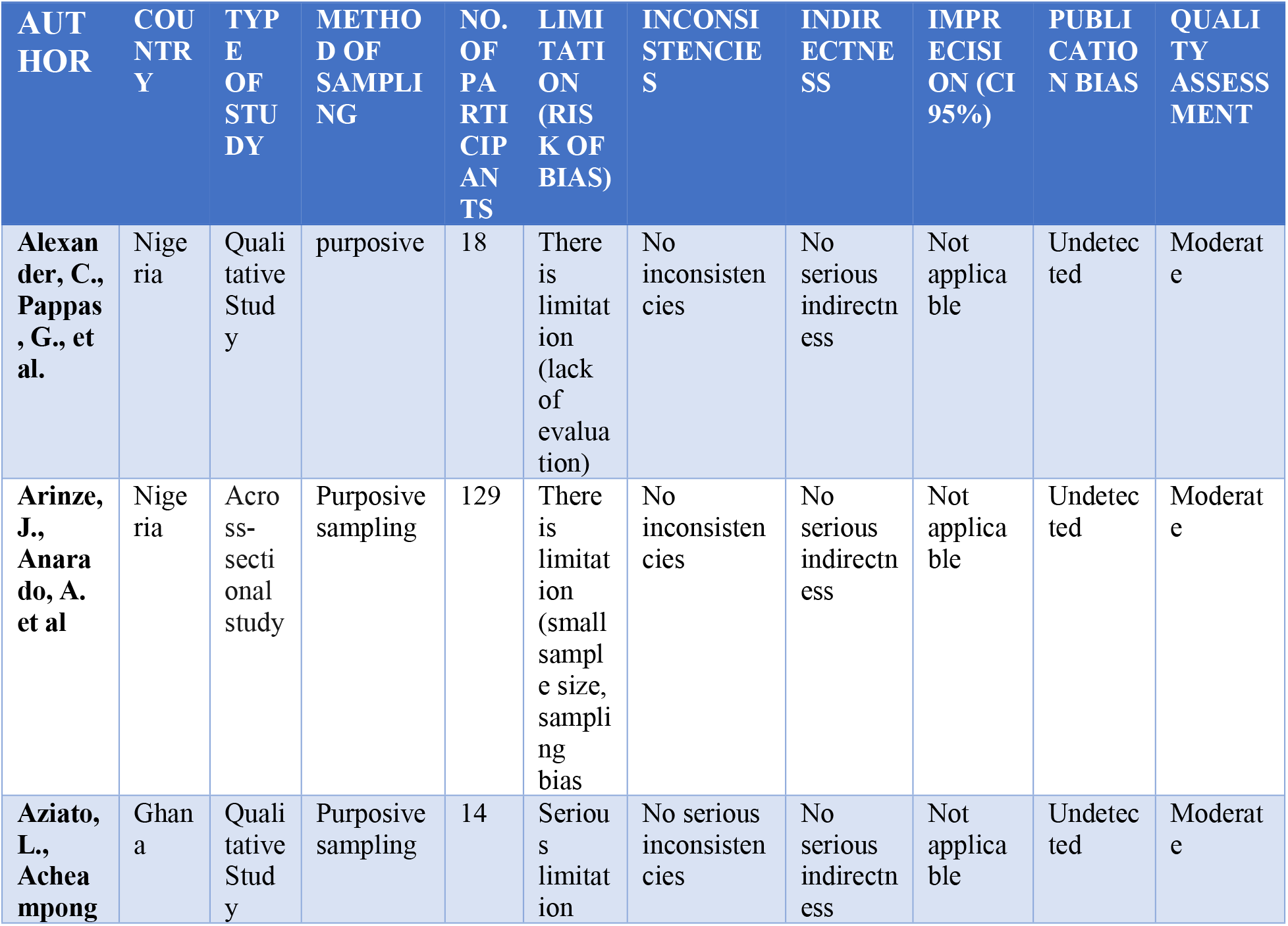
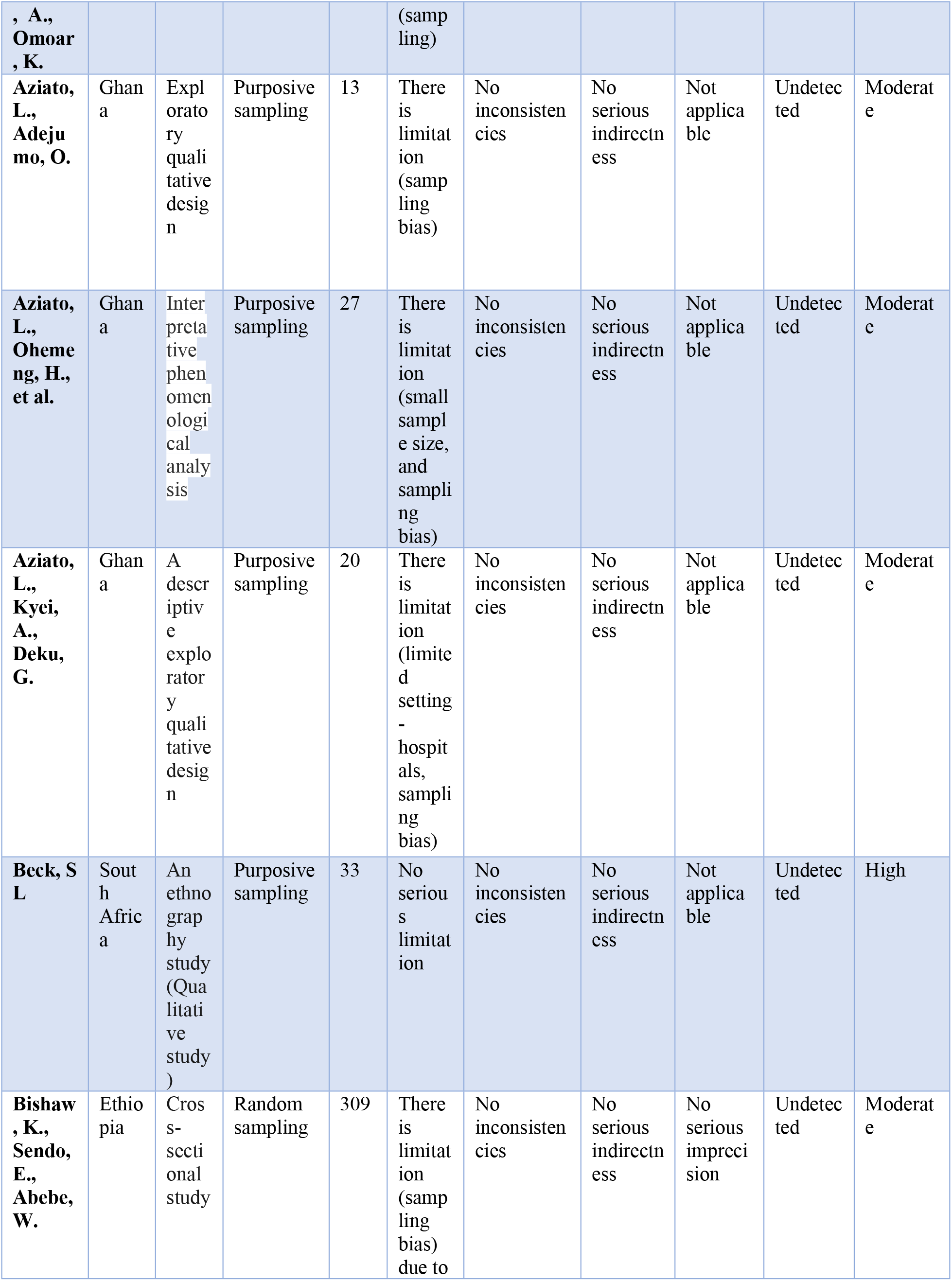
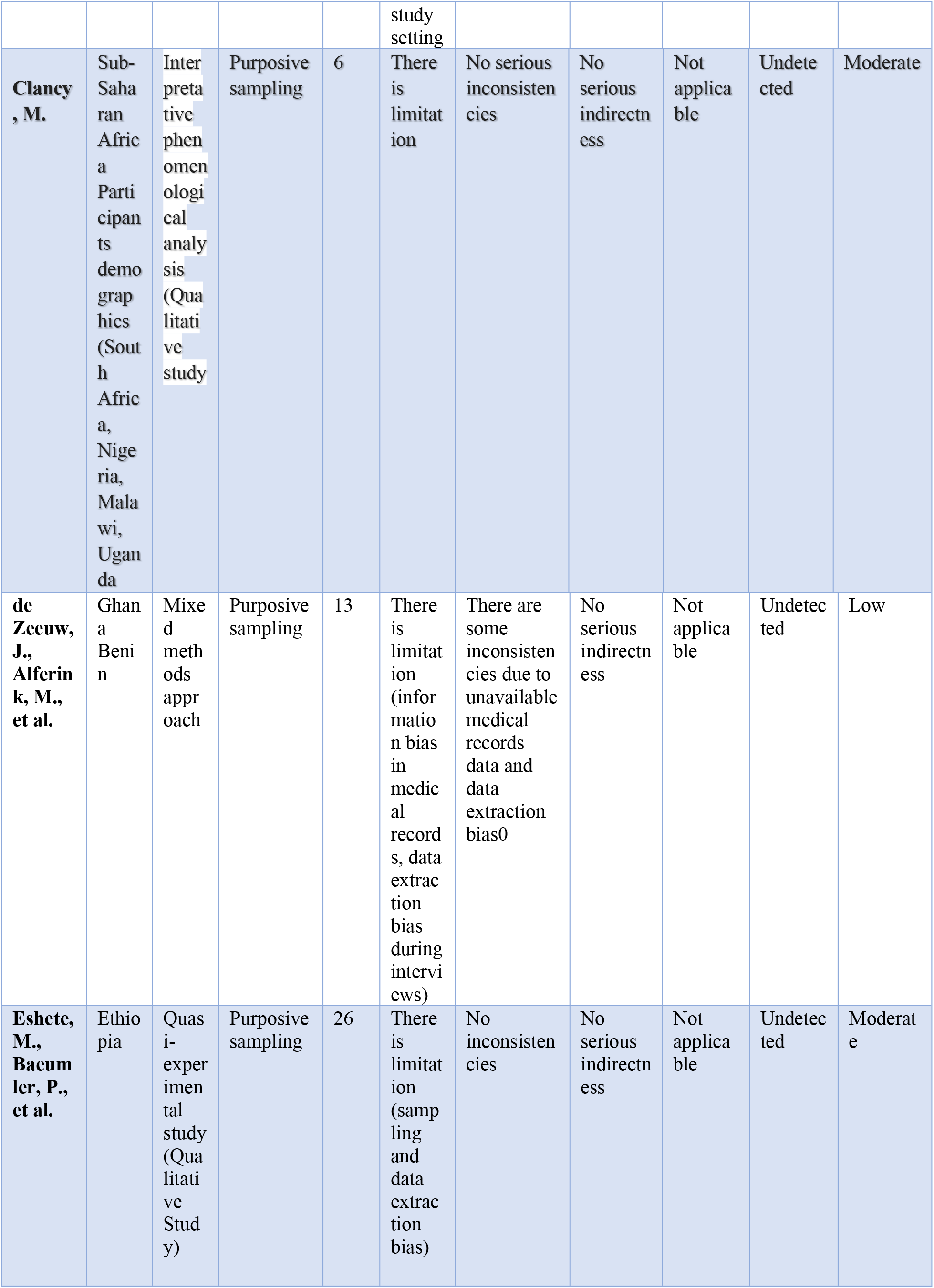
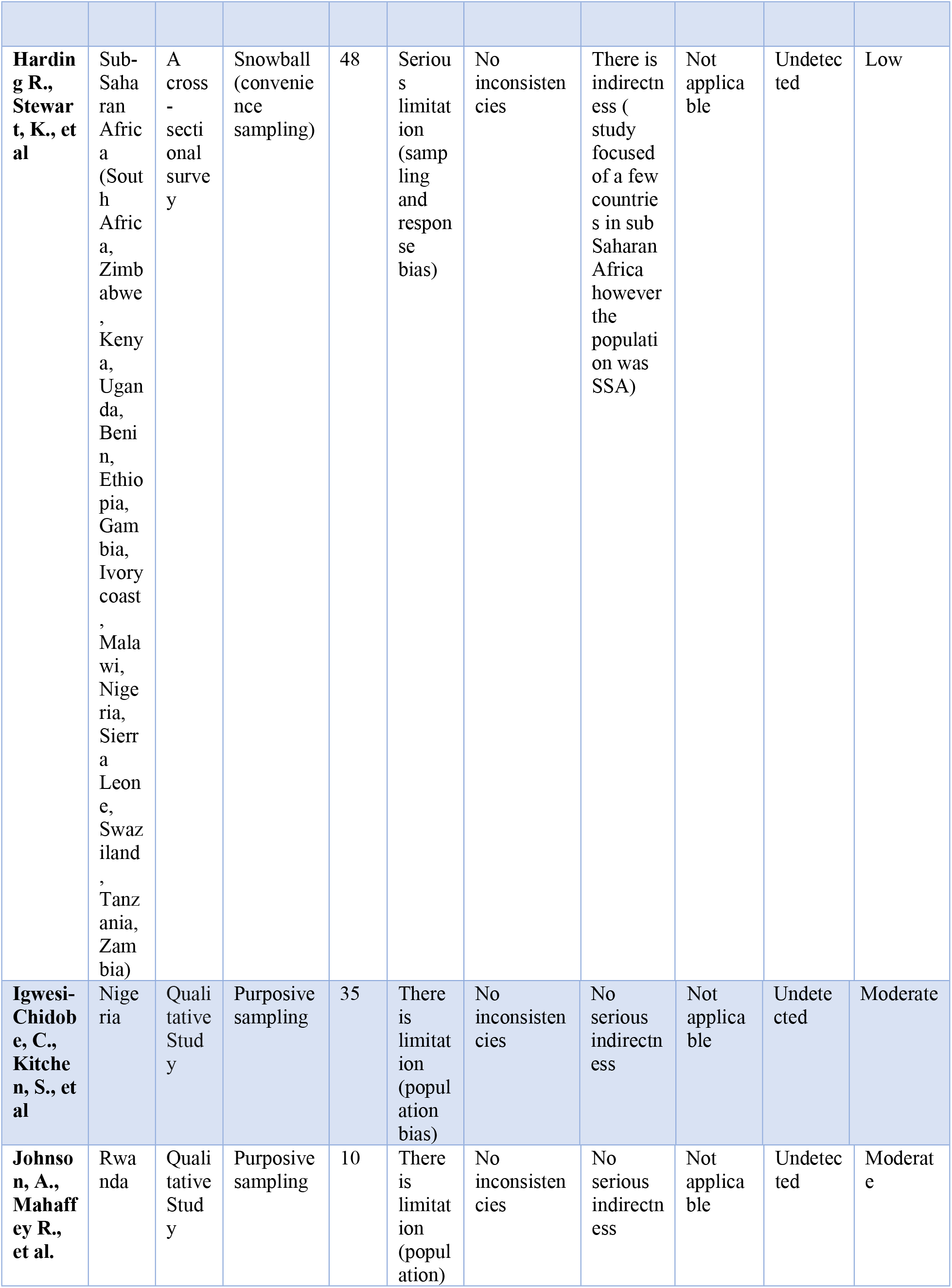
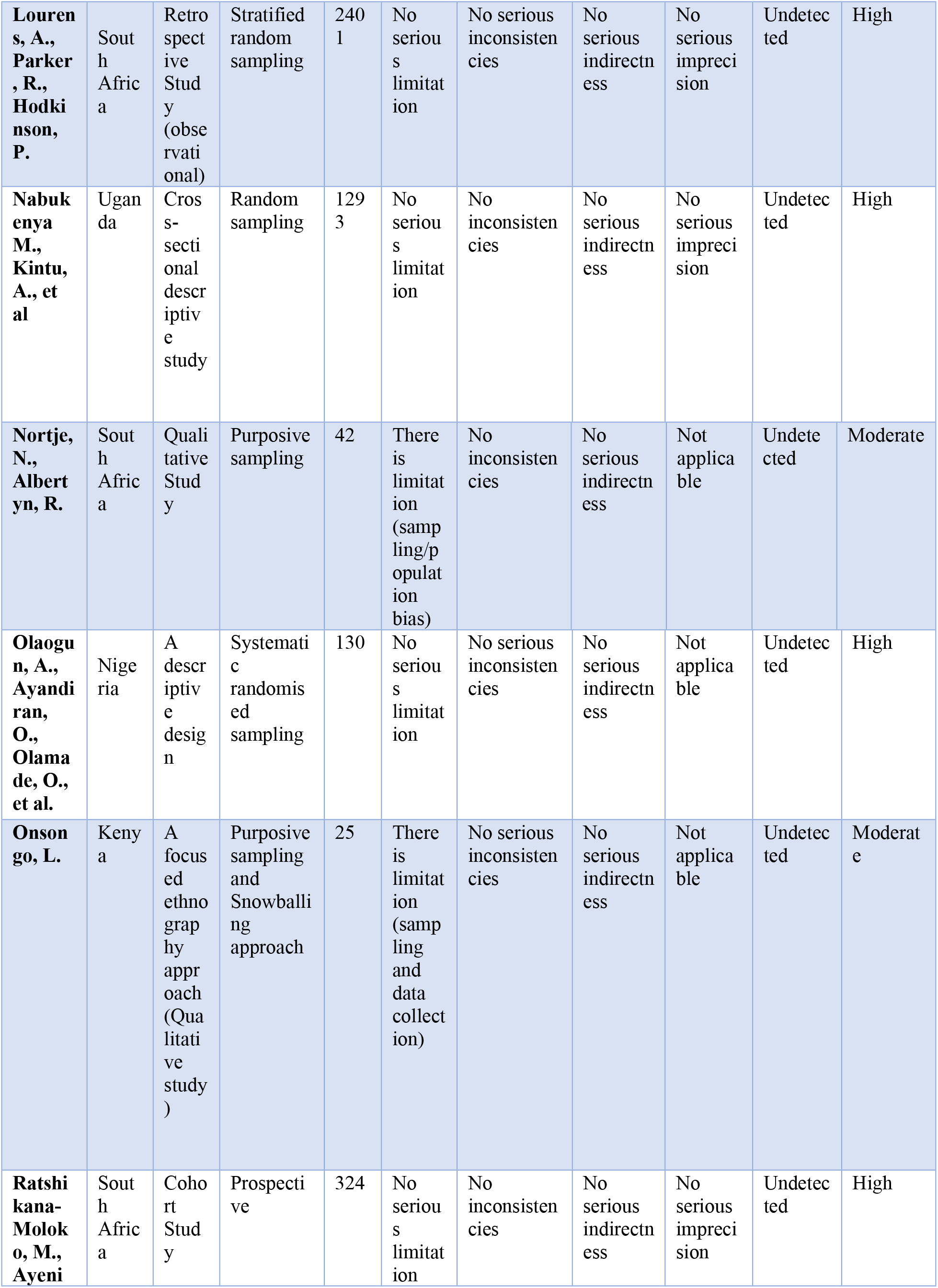
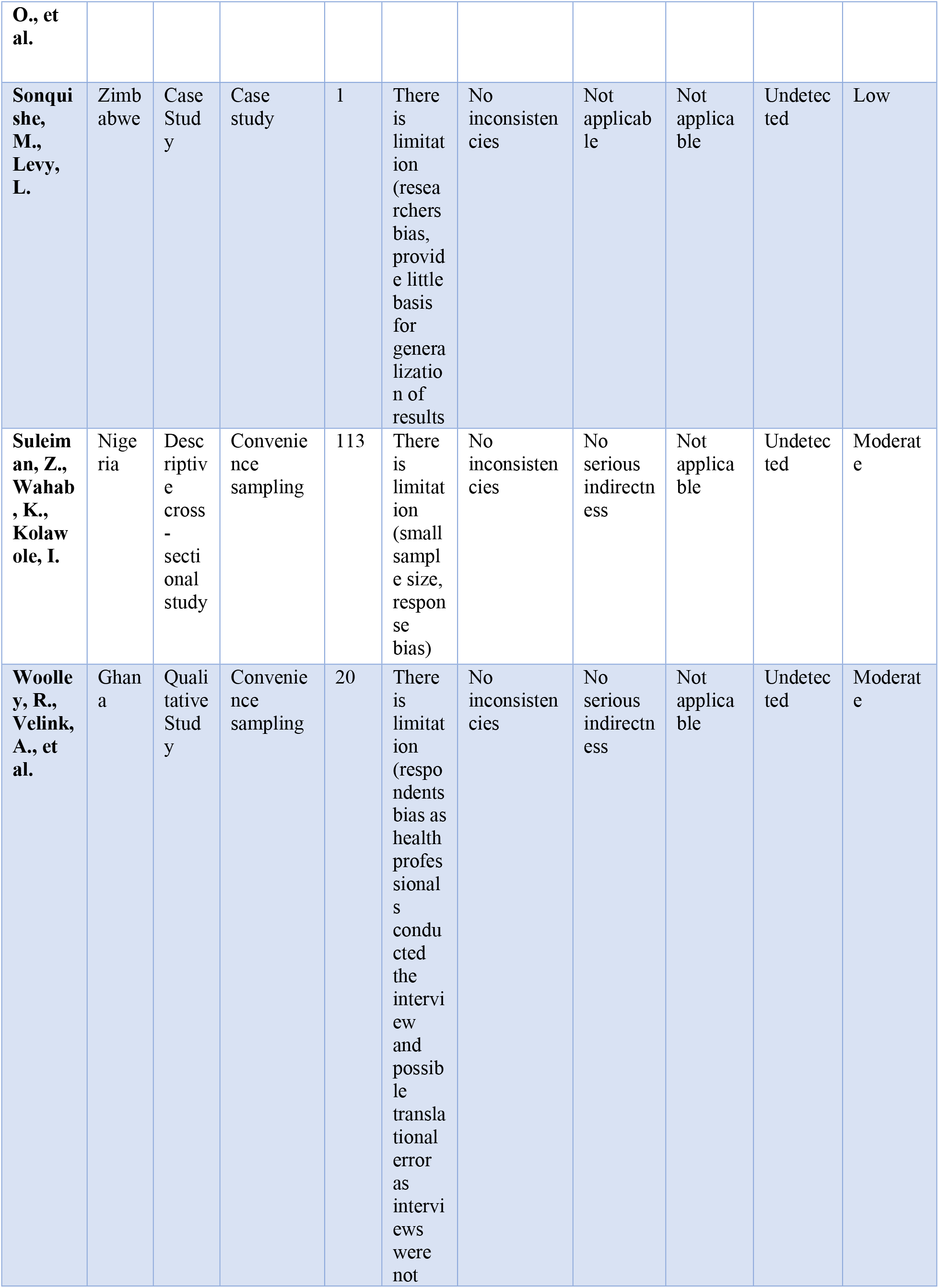
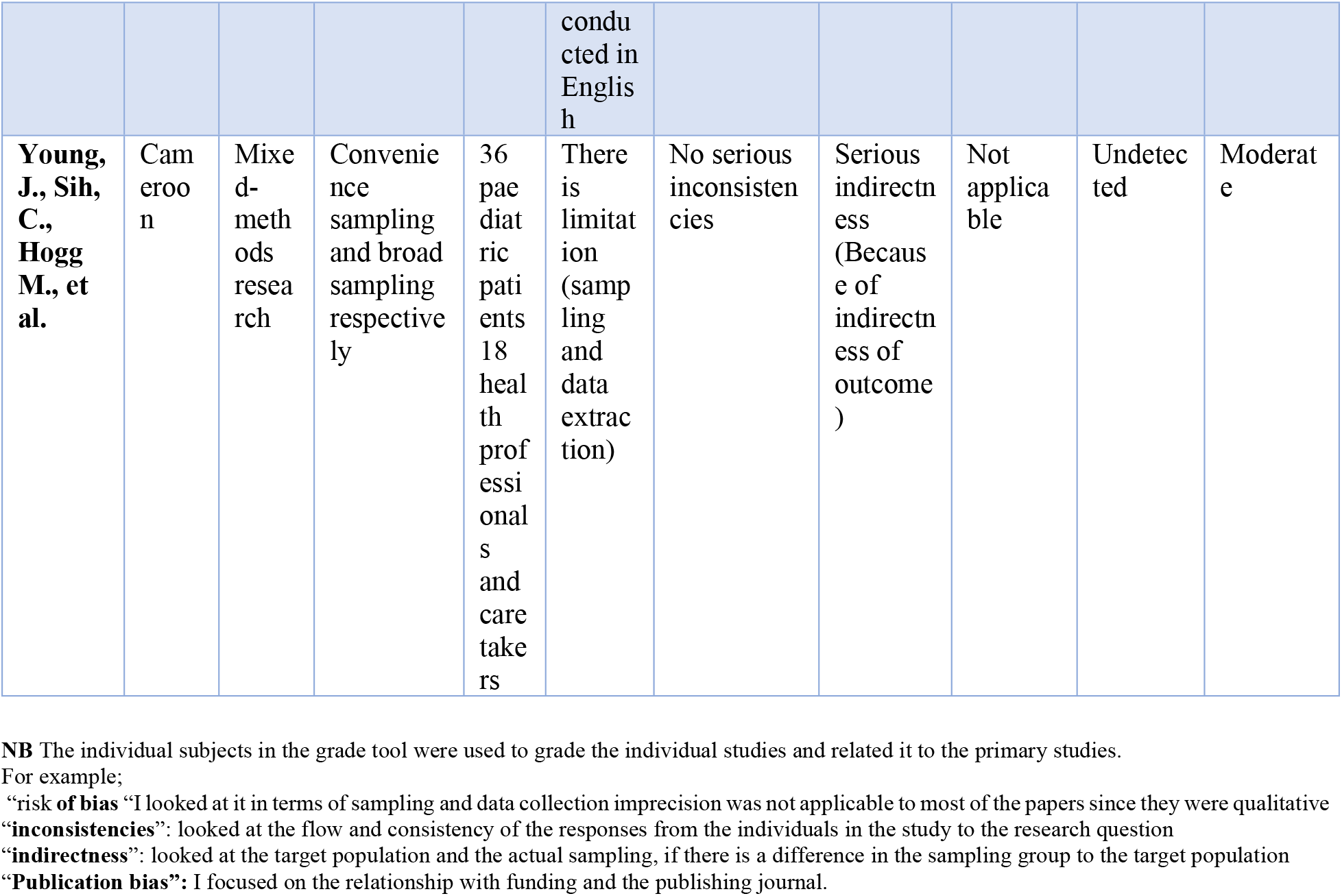
Characteristics And Quality Assessment of the included articles using The Grading of Recommendations Assessment, Development, and Evaluation (Grade)

### Study population

Examination of the 24 studies included in this systematic review identified that five studies were conducted in Nigeria (Alexander et al.,2015, Arinze et al., 2018, Igwesi-Chidobe et al., 2017, Olaogun et al., 2008, Suleiman et al., 2016), a further five studies were from Ghana (Aziato et al., 2017, Aziato & Adejumo, 2014, Aziato et al. 2016, Aziato et al., 2017, Woolley et al., 2016), four from South Africa (Beck, 2000, Lourens et al., 2020, Nortje & Albertyn, 2015, Ratshikana-Moloko et al., 2020), two from studies were from Ethiopia (Bishaw et al., 2020, Eshete et al., 2019), two from SSA (Clancy, 2014, Harding, 2003), and one study from from Ghana & Benin (de Zeeuw et al., 2015), Rwanda (Johnson et al., 2015), Uganda (Nabukenya et al., 2015), Kenya (Onsongo, 2020), Zimbabwe (Sonquishe & Levy, 1990), and Cameroon (Young et al., 2018).

### Narrative thematic Analysis

Four themes emerged from the included literature, which included 8 articles focusing on religious/spiritual and inherited beliefs, with a further 7 articles focusing on limited knowledge and health literacy. The third theme focused on resource constraints which included 5 articles and the final theme on cultural and societal norms which included 4 articles.

### Religion, Spiritual and Inherited Beliefs

Eight studies were included in this theme (Ratshikana-Moloko, et al., 2020, Aziato & Adejumo, 2014, Sonqishe & Levy,1990, Nortjé & Albertyn, 2015, Beck 2000, Igwesi-Chidobe, et al., 2017, Aziato, et al., 2016, Aziato, et al., 2017), of which two sub-themes emerged which were pain as a source of punishment/spiritual forces/bewitchment and pain as a source of natural phenomena.

#### Punishment/spiritual forces/bewitchment

Six studies were included in this subtheme; (Ratshikana-Moloko, et al., 2020, Aziato & Adejumo, 2014, Sonqishe & Levy 1990, Nortjé & Albertyn, 2015, Beck 2000, Igwesi-Chidobe, et al., 2017). Evidence emerged suggests that patients believed their source of illness were a result of offending God, therefore, seeking a closer connection with God and asking forgiveness for sins was the best remedy (Aziato & Adejumo, 2014). Patients were less likely to complain of pain or take pain medication when receiving spiritual care than those who did not receive spiritual care, therefore had exacerbated pain and requirement for more pain medication (Ratshikana-Moloko, et al., 2020). Further evidence identified that surgical patients and relatives believed God would see them through surgery successfully resulting in subsidized pain, therefore, depended on God for healing (Aziato & Adejumo, 2014). In Zimbabwe, ancestral spirit known as “Vadzimu” is the cause of diseases and inflicts pain to individuals who do not please them. People therefore seek traditional help to appease “Vadzimu” to be safe from such sufferings/pains (Sonqishe & Levy 1990). This concept is also highlighted by Beck (2000) and identified that, cancer and its associated pain is not a disease like virus such as flu however, it is a punishment from the ancestors for not living a good life or offending them (Nortjé & Albertyn, 2015). Evidence also suggests that close neighbours, relatives and friends bewitch individuals by causing them pain (Igwesi-Chidobe, et al., 2017).

#### Pain as a source of natural phenomena

Conversely, two studies were included in this sub-theme; (Aziato, et al., 2016, Aziato, et al., 2017). Evidence reported pain as natural and instituted by God (Aziato, et al., 2016, Aziato, et al., 2017). It is therefore normal for women undergoing childbirth to experience pain, successfully enduring the pain symbolizes womanhood and strength. Women that expressed pain and requested pain relieve were termed as weak and a disgrace. Therefore, women should feel proud to undergo labour pain without expressing it (Aziato, et al., 2016; Aziato, et al., 2017). Due to these societal expectations, women look forward to undergoing childbirth without expressing pain as it is rewarding.

### Limited knowledge and health literacy

Literatures examined in this theme explored limited knowledge and health literacy and the resulting impact on pain management approaches. A total of seven studies were included in this theme; (Arinze, et al 2018, Lourens, et al., 2020, Johnson, et al., 2015, Eshete, et al. 2019, Olaogun et al, 2008, Nabukenya, et al, 2015, Suleiman, et al., 2016). Nabukenya et al., (2015), conducted a cross-sectional descriptive study to determine women’s attitude and knowledge on use of labour analgesics in Uganda. The study indicated that, 93% of the participants had no knowledge on analgesia used during labour and only 7% had knowledge of labour analgesia. Findings indicated that, mothers primarily reason for not taking labour analgesics was to experience natural childbirth (45%), others were it being against the will of God, baby may be affected, may lead to C/S etc. Evidence suggests women preferred to go through labour without any pharmacological intervention due to these misconception and myths associated with pain medication and chose natural childbirth resulting labour pain. Nonetheless, Bishaw et al., 2020, examined obstetric care givers knowledge and use of labour pain relief methods in Ethiopia. Results indicated that, 93% of the participants knew about pain relief methods. However, only 15% of participants knew about pharmacological relief methods. Similar findings were identified by Arinze, et al., 2018, among Nigerian women, on their perception on labour pain. Results indicated that women during labour had limited knowledge on the various pain relief methods. Majority (61%) of women knew psychotherapy (breathing exercises, massages etc.) pain relief methods (Eshete, et al., 2019; Bishaw et al., 2020). Conversely indifferent among health professionals, the most frequently used pain relief method was psychotherapy due to limited knowledge on pharmacological approaches (Lourens, et al., 2020).

The difference in findings from Nabukenya et al., (2015) and Bishaw et al., 2020 could be a results of sample size. Nabukenya et al., (2015), had a larger sample size of 1,293 compared to Bishaw et al., 2020 sample size of 299 of which according to Zamboni, J., (2021), large sample size provides more reliable results with greater precision. Another reason for difference in findings could be the characteristic of study population. The study population of Bishaw et al., 2020, were health professionals who are likely to be more informed or knowledgeable on the study area to that of Nabukenya et al., (2015), who’s study population were women in general.

A retrospective study conducted by Lourens, et al., (2020) on prehospital acute traumatic pain assessment and management practices indicated that, 97% of patients were experiencing pain, however, only 3% received any form of analgesia. 18% of the total population had pain score recorded and the study indicated an association among patients who had pain score to receiving analgesia, however, the strength of association was week. According to Lourens, et al., (2020), a barrier commonly highlighted for the constraint to pain assessment and management was knowledge deficit, attributed to limited attention to pain assessment and management during initial training and a lack of ongoing education.

As well as health professionals limited knowledge, patients also were found to lack knowledge. A study conducted by Olaogun, et al., (2008), indicated that, mothers believed infants below two-month-old do not experience pain. This supports the common myth in SSA that neonates do not experience pain, do not remember painful experiences and when they experience pain, the pain does not have any negative effect on them (Byers & Thornley, 2004).

### Resource Constraint

Five studies were included in this theme, which explored resource constraint; (Aziato, et al., 2017, Bishaw, et al., 2020, Harding, et al., 2003, Onsong 2020, Clancy, 2014). The above studies evidence indicated that, the lack of resources and limited availability of equipment and tools as well as inaccessibility of pain relief medication were found to be significant barriers to the treatment of pain. Organizational factors were also found to be a barrier to pain management resources (Harding, et al.,2003). Evidence suggested that there were challenges to providing pain relief, which included unavailability of some pain medication especially opioids due to government restrictions, funders restrictions as well as prohibitive drug cost (Onsongo, 2020; Suleiman, et al., 2016). In some SSA countries, the ratio of midwives/nurses to patients are very low leading to increase workload on midwives (Aziato, et al., 2017; Bishaw, et al., 2020). Findings indicated that health professionals due to the workload are tasked-oriented thus they only focus on duty to be performed than looking at other aspect of nursing care which pain management is inclusive (Clancy (2014).

### Cultural and Societal Norms

Four studies were included in this theme, which is cultural and societal norms; (Young, et al., 2018, Woolley, et al., 2016, de Zeeuw, et al., 2015, Alexander, et al., 2015). The evidence suggests that society plays a major role on an individual’s expression of pain. In Nigeria, some cultures believe in a philosophy of non-expression of pain to demonstrate bravery and disapprove on those who express pain externally. According to Woolley, et al., (2016), patients do not express pain during treatment due to the fear of being shouted at by nurses/health professionals. Most health professionals believe that patients exaggerate pain (de Zeeuw, 2015). These findings indicates that, since health professionals are part of the society in which these are the accepted way of expressing pain, believe patients who express pain overtly may exaggerate their pain experience.

The role of society’s influence on pain expression is particularly apparent regarding gender. According to Young, et al., (2018), male children are more stoic towards pain compared to female children. Male children refused to acknowledge pain in the presence of their parents through non-verbal cues (eye downcast, avoiding eye contact and fidgeting). Some also asked their parents to answer questions concerning their pain. These findings identify those societal expectations of mothers on pain influences imposed on their children. Men who express pain are termed as weak and male children referred to as “girls” or “a baby” when they express pain (Alexander, et al., 2015). Expressions of pain are not acceptable, and children learn this from an early stage.

## Discussion

This study identified the sociocultural barriers to effective pain management from the perspective of different subcultures. It also explored patients and Health workers perspective on sociocultural barriers to effective pain management approaches in SSA. Healthcare professionals should be aware of how the society and cultural beliefs of their patients influence their expression of pain. Under-treatment or over-treatment might occur in pain management if health workers are not aware of the cultural norms associated with pain and pain expression, as experience of pain is both personal and subjective.

A key objective of this systematic review was to determine the prevalence of pain among people living in SSA. Globally, the prevalence, seriousness, disparities, and vulnerability to pain varies across nations and sub-regions. In SSA and many parts of the developing world the prevalence and severity of pain is often much higher resulting from limited capacity to effectively diagnose pain coupled with unavailability of efficacious treatment options (Miftah *et al*., 2017). Indeed Miftah *et al*. (2017) have noted that the prevalence and severity of pain especially in West Africa is amplified by lack of access to health facilities, late presentation, inadequate diagnosis, and treatment unavailability. In addition, these factors in developing countries have been compounded by disproportionate distribution of global burden of many diseases and their concomitant outcomes including pain (Hammond *et al*., 2015). In fact, diseases that no longer present health threats at global level such as sickle cell, polio, malaria, HIV/AIDS and other many infectious diseases are still significant source of pain and mortality in SSA (Noah and Fidas, 2000). The harsh impact of these inequalities is that, for most SSA countries there is very limited access to healthcare and low disease response capacity making individuals including children in these countries much more vulnerable to disease, infection, and pain than other regions globally. Combined with these factors there is a requirement to take account of high populations, poverty, illiteracy issues, political instability, unavailability of pain medication, belief system impact on pain access and management in SSA is often problematic. According to Clancy (2014), attitudinal barriers also impact greatly as cultural practices, customs and beliefs all influence how pain assessment and management is conducted and how individuals express pain. Key findings from this systematic review demonstrates that there are variances in the barriers leading to effective and efficient pain prevalence in SSA, which include the sociocultural barriers to successful pain management. In addition, results revealed several socio-cultural factors; Religion, Spiritual and Inherited Beliefs, Limited Knowledge Literacy, Resource Constraint and Cultural and Societal Norms contribute to imperfect pain management in SSA. The prevalence of frequent pain experienced in SSA demonstrated gender differences predominantly as the evidence meeting the eligibility criteria for this systemic review were investigating labour and childbirth pain, palliative pain, acute and chronic medical conditions, and paediatric pain.

Another key objective of this systematic review was exploring the socio-cultural factors which influence accessing and management of pain in SSA. According to Albertyn et al (2009), there is limited evidence available focusing on the influence of society and culture on pain. According to Aziato et al (2015), pain response is influenced by the society and cultural background of the individual. This factor is also identified by Ufashingabire et al (2016) and suggest that cultural patterns significantly control or manipulate individuals in all their dimensions of life including how pain is expressed. The responses to a painful stimulus are not an innate characteristic but rather cultivated through socialization from the individual’s culture (Aziato et al., 2015). This is suggestive that culture and society, have significant impact on individual behaviour which encompasses beliefs, knowledge, practices, values, and everyday life activities (Callister 2003). These characteristics conclude that the expression of behavioural signs both verbal and nonverbal depends largely on the sociocultural exposure of the individual experiencing pain.

The findings from this systematic review provide a clear understanding on the concerns that sociocultural influences have effect on pain management approaches in SSA. The evidence indicates that societal and cultural factors impact to pain management and linked with spirituality and religion beliefs. The conviction that pains is a natural phenomenon and should be experienced during labor with biblical doctrine among the female population and health professionals suggestive that God could help through uncomplicated labour and safe delivery alone (Aziato et al., 2016). Thus, to relieve labour pain, women need to connect with God, and people going through pain prayed and requested prayer support from their spiritual leaders. These findings are consistent to other studies where religious rituals, namely blessed oil, and holy water, use of relics of saints, holy icons, offering names for pleas and pilgrimage were used to relieve pain and promote healing (Fouka, et al., 2012) (Kaphle et al., 2013). Pregnancy and labour pain were as well associated with evil spirits with enduring pain viewed as a form of punishment/spiritual force/bewitchment (Ratshikana-Moloko, et al.,2020). Nortjé and Albertyn (2015) suggest that stories and folklore are abundant and important in some cultural group, to them pain is often identified as an outcome of defiant behaviour by the antagonist. Individuals therefore have a preconceived perception that, pain is a form of punishment for an offence against an ancestor. Therefore, pain can only be controlled when the ancestors are appeased.

Lack of health literacy is also a factor to ineffective pain management in SSA. The evidence indicates that due to the lack of knowledge, health professionals have regarding analgesia as well as effective pain approaches significantly influence access and effective pain management. Those that were enlightened about pain medication had myths and misinformation on pain and pain management approaches. Evidence suggests that most health professionals are not taught pain medication/ analgesia during their training programme as well as their practical exposure (Taneja et al., 2004). A study conducted by Lohman et al (2010) showed that, a significant number of health professionals in Africa report inadequate knowledge on palliative care and pain management. Cleary et al (2013), also indicated that, some are not allowed to prescribe pain medications (oral morphine). In Nigeria majority of health practitioners had received no formal education on pain management (Cleary et al 2013). This was confirmed in a study conducted by Low et al (2018), that there is a major gap in knowledge on palliative care and pain management among African physicians. The fear of health professionals not to give some pain medication is due to the lack of understanding of the treatment value of controlled medicines and according to Ufashingabire et al (2016), this can be primarily tackled by the education and experience of health practitioners.

Research findings examining organizational restrictions such as government policies is another sociocultural barrier impacting on effective pain management. According to Anderson (2010), most countries in SSA reported severe restrictions in accessing pain medications especially morphine. These restrictions were primarily due to funding, lack of adequate distribution systems and widespread fear of addiction (Israels et al 2010). Evidence suggests that policies restricting access and dispensing of analgesics are due to limited appropriate prescribing professionals among SSA. (Al-Shamsi 2017). According to Nchako et al (2018), the current restrictive laws in SSA, are primarily aimed at preventing drug diversion or abuse due to perceived fear of addiction, and that the rules to obtaining licenses are so stringent that most pharmacies do not stock analgesics specifically opioids. A study conducted in Nigeria and Cameroon revealed that, some countries had just one government pharmacy each that stocks oral morphine, which makes access to these drugs very limited (Anderson 2010). In addition, linked with the phenomenon of addiction, health professionals are inclined to abstain from using opioids when it is needed to be administered although several studies have indicated that addiction or psychological dependence when opioids are used is rare in terminal and life-limiting illnesses (Nchako et al 2018). Coupled with this are human resource/ infrastructure and financial limitation to effective pain management. According to Lee and Akhtar (2011), heavy workload as well as the patient-to-nurse ratio are low regardless of the number of patients or the acuity of patients on the unit and this consequently leads to work burnout. This is similar to the findings from Toh et al., (2012), who indicated that workload, lack of debriefing sessions, and lack of career progression may contribute to burnout, job dissatisfaction and stress leading to less focus on pain management.

Culture and societal norms are considered sociocultural factors to effective pain management after analysing the papers included in the studies. The evidence indicates that culture within given societies largely has an influence on population behaviours which takes account of societal beliefs, knowledge, practices, values, and everyday life activities and this consequently affect how an individual is expected to express or endure pain. In some part of Sub-Saharan Africa, it is expected of an individual to endure pain, to keep calm and not let their surroundings know they are in pain Callister (2003). The expression of pain among individuals is perceived as a weakness on the victim and there is the believe that remaining calm and focused when an individual is experiencing pain helps to alleviate the pain experienced (Ufashingabire et al., 2016). Evidence suggests that when a woman cry/moan when in pain especially during delivery, they will remain same throughout all their deliveries (Aziato et al 2015). Another study by Finnstrom et al (2006) indicated, women during childbirth are not expected to cry/wail as it was seen as unacceptable ways of expressing pain. It is believed that life is made up of sufferings and pain and involves self-control and tolerance, this emphasis on the value of endurance and glory/ reward from others when they showed courage (Keynan 2001). Meaning a reward awaits you when you endure pain and don’t show it.

The evidence suggests that there are gender differences linked with culture in SSA. In SSA cultural factors such as gender influence pain expression and management as, men in SSA are not supposed to cry or show any sign of pain as this expression is perceived as a sign of weakness, and cultural influences expectations regarded to endure pain. In SSA culture, it is a taboo for a man to express pain, this tradition is instilled into children early in childhood not to show or express pain when in pain or when they hear sad news (Keynan 2001; Nortjé & Albertyn 2015; Young et al 2018). A further consideration in the cultural influences in effective pain management is that health care practitioners are also impacted by cultural factors in their practice in SSA. Due to the socio-cultural influencer’s health professionals perceive patient’s expression of pain as an exaggeration of their pain and was therefore the expectation of patients to endure pain (de Zeeuw et al 2015; Woolley, et al., 2016).

## 3 Conclusion

The evidence presented in this systematic review examining the factors influencing effective pain management in SSA is at the factors such as sociocultural influences are under explored particularly in low-middle income countries (LMICs). In addition, LMIC have perceived ideology with regards to the causes of diseases and pain due to their cultural context. The findings suggest that significance of pain in the SSA culture is seen as a natural part of disease or injury and not as is the case in Western medicine as a separate entity. The results suggest the expression or acknowledgement of pain can almost be described as a cultural taboo associated with weakness and a lack of honour and courage amongst gender. Folklore describing the stories of ancestors are often used to train African children to deal with pain in a stoical and resilient manner. This greatly influences the reaction of members of these cultures towards pain, which explains why health care providers need to show greater sensitivity to the cultural expression of pain. Coupled with these are the lack of education, resources, and governmental restrictions to pain dispensing protocols. This systematic review indicates that, pain assessment and management particularly in SSA is under-recognised because of the sociocultural influences. Findings confirm the need to scale up scientific research aimed at proposing a culturally context standardised procedure and methods that could be adopted for pain assessment and management among people living in Sub-Saharan Africa. This will contribute a great effectively assessing and managing in among these group.

## Data Availability

All relevant data are within the manuscript and its Supporting Information files.

## Declarations

### Ethics approval

The study was registered with the Academic Ethics Committee for Bangor University and was granted exemption from requiring ethics approval.

## References

1. Al-Shamsi M. Addressing the physicians’ shortage in developing countries by accelerating and reforming the medical education: Is it possible?. Journal of Advances in Medical Education & Professionalism. 2017 Oct;5(4):210.

2. Albertyn R, Rode H, Millar AJ, Thomas J. Challenges associated with paediatric pain management in Sub Saharan Africa. International Journal of Surgery. 2009 Jan 1;7(2):91–3.

3. Alexander CS, Pappas G, Henley Y, Kaiza Kangalawe A, Olusegun Oyebola F, Obiefune M, et al. Pain Management for Persons Living With HIV Disease: Experience With Interprofessional Education in Nigeria. Am J Hosp Palliat Med [Internet]. 2015 Aug;32(5):555–62.Available from: http://ezproxy.bangor.ac.uk/login?url=http://search.ebscohost.com/login.aspx?direct=true&db=rzh&AN=109819042&site=ehost-live

4. Anderson C. Presenting and evaluating qualitative research. American journal of pharmaceutical education. 2010 Oct 1;74(8).

5. Arinze JC, Anarado AN, Nwonu EI, Ogbonnaya CE, Maduakolam IO, Arinze OC. Perceptions of labour pain among gravid women in Enugu, Nigeria. African J Midwifery Women’s Heal [Internet]. 2018 Oct;12(4):194–9. Available from: http://ezproxy.bangor.ac.uk/login?url=http://search.ebscohost.com/login.aspx?direct=true&db=rzh&AN=133011320&site=ehost-live

6. Aziato L, Acheampong AK, Umoar KL. Labour pain experiences and perceptions: a qualitative study among post-partum women in Ghana. BMC Pregnancy Childbirth [Internet]. 2017 Feb 22;17:1–9. Available from: http://ezproxy.bangor.ac.uk/login?url=http://search.ebscohost.com/login.aspx?direct=true&db=rzh&AN=121474065&site=ehost-live

7. Aziato L, Adejumo O. PSYCHOSOCIAL FACTORS INFLUENCING GHANAIAN FAMILY CAREGIVERS IN THE POST-OPERATIVE CARE OF THEIR HOSPITALISED PATIENTS. Africa J Nurs Midwifery [Internet]. 2014 Sep;16(2):112–24. Available from: http://ezproxy.bangor.ac.uk/login?url=http://search.ebscohost.com/login.aspx?direct=true&db=rzh&AN=115965744&site=ehost-live

8. Aziato L, Kyei AA, Deku G. Experiences of midwives on pharmacological and non-pharmacological labour pain management in Ghana. Reprod Health. 2017;14(1):1–8.

9. Aziato L, Ohemeng HA, Omenyo CN. Experiences and perceptions of Ghanaian midwives on labour pain and religious beliefs and practices influencing their care of women in labour. Reprod Health [Internet]. 2016 Nov 14;13:1–7. Available from: http://ezproxy.bangor.ac.uk/login?url=http://search.ebscohost.com/login.aspx?direct=true&db=rzh&AN=119486566&site=ehost-live

10. Beck SL. An ethnographic study of factors influencing cancer pain management in South Africa. Cancer Nurs [Internet]. 2000;23(2):91–100. Available from: http://ezproxy.bangor.ac.uk/login?url=http://search.ebscohost.com/login.aspx?direct=true&db=rzh&AN=107112161&site=ehost-live

11. Bishaw KA, Sendo EG, Abebe WS. Knowledge, and use of labour pain relief methods and associated factors among obstetric caregivers at public health centers of East Gojjam zone, Amhara region, Ethiopia: a facility based cross-sectional study. BMC Pregnancy Childbirth [Internet]. 2020 Mar 23;20(1):1–9. Available from: http://ezproxy.bangor.ac.uk/login?url=http://search.ebscohost.com/login.aspx?direct=true&db=rzh&AN=142383492&site=ehost-live

12. Burton EF, Suen SY, Walker JL, Bruehl S, Peterlin BL, Tompkins DA, Buenaver LF, Edwards RR, Campbell CM. Ethnic differences in the effects of naloxone on sustained evoked pain: A preliminary study. Diversity and equality in health and care. 2017;14(5):236.

13. Callister LC. Cultural influences on pain perceptions and behaviors. Home Health Care Management & Practice. 2003 Apr;15(3):207–11.

14. Calvillo ER, Flaskerud JH. Evaluation of the pain response by Mexican American and Anglo American women and their nurses. Journal of Advanced Nursing. 1993 Mar;18(3):451–9.

15. Clancy MA. Difficulty, despair and hope – an insight into the world of the health professionals treating paediatric pain in sub-Saharan Africa. J Res Nurs [Internet]. 2014 May;19(3):191–210. Available from: http://ezproxy.bangor.ac.uk/login?url=http://search.ebscohost.com/login.aspx?direct=true&db=rzh&AN=95617606&site=ehost-live

16. Cleary J, Powell RA, Munene G, Mwangi-Powell FN, Luyirika E, Kiyange F, Merriman A, Scholten W, Radbruch L, Torode J, Cherny NI. Formulary availability and regulatory barriers to accessibility of opioids for cancer pain in Africa: a report from the Global Opioid Policy Initiative (GOPI). Annals of Oncology. 2013 Dec 1;24:xi14–23.

17. de Zeeuw J, Alferink M, Barogui YT, Sopoh G, Phillips RO, van der Werf TS, et al. Assessment and Treatment of Pain during Treatment of Buruli Ulcer. PLoS Negl Trop Dis. 2015;9(9):1–10.

18. Eshete MT, Baeumler PI, Siebeck M, Tesfaye M, Wonde D, Haileamlak A, et al. The views of patients, healthcare professionals and hospital officials on barriers to and facilitators of quality pain management in Ethiopian hospitals: A qualitative study. PLoS One. 2019;14(3):1–20.

19. Finnström B, Söderhamn O. Conceptions of pain among Somali women. J Adv Nurs [Internet]. 2006 May 15;54(4):418–25. Available from: http://ezproxy.bangor.ac.uk/login?url=http://search.ebscohost.com/login.aspx?direct=true&db=rzh&AN=20656764&site=ehost-live

20. Fouka G, Plakas S, Taket A, Boudioni M, Dandoulakis M. Health-related religious rituals of the G reek O rthodox C hurch: their uptake and meanings. Journal of Nursing Management. 2012 Dec;20(8):1058–68.

21. Guyatt GH, Oxman AD, Schünemann HJ, Tugwell P, Knottnerus A. GRADE guidelines: a new series of articles in the Journal of Clinical Epidemiology. Journal of clinical epidemiology. 2011 Apr 1;64(4):380–2.

22. Hammond CK, Dogbe J, Paintsil V, Tutu L, Owusu M. Pain description and presentation in children admitted to a teaching hospital in Ghana. J Cancer Prev Curr Res. 2015;2(4):00046.

23. Harding R, Stewart K, Marconi K, O’Neill JF, Higginson IJ. Current HIV/AIDS end-of-life care in sub-Saharan Africa: a survey of models, services, challenges and priorities. BMC Public Health [Internet]. 2003;3:33. Available from: http://ovidsp.ovid.com/ovidweb.cgi?T=JS&PAGE=reference&D=med5&NEWS=N&AN=14572317

24. Herr K, Coyne PJ, McCaffery M, Manworren R, Merkel S. Pain assessment in the patient unable to self-report: position statement with clinical practice recommendations. Pain management nursing. 2011 Dec 1;12(4):230–50.

25. Igwesi-Chidobe CN, Kitchen S, Sorinola IO, Godfrey EL. “A life of living death”: the experiences of people living with chronic low back pain in rural Nigeria. Disabil Rehabil [Internet]. 2017 Apr 15;39(9):779–90. Available from: http://ezproxy.bangor.ac.uk/login?url=http://search.ebscohost.com/login.aspx?direct=true&db=rzh&AN=139413418&site=ehost-live

26. Israels T, Ribeiro RC, Molyneux EM. Strategies to improve care for children with cancer in Sub-Saharan Africa. European journal of cancer. 2010 Jul 1;46(11):1960–6.

27. Johnson AP, Mahaffey R, Egan R, Twagirumugabe T, Parlow JL. Perspectives, perceptions and experiences in postoperative pain management in developing countries: A focus group study conducted in Rwanda. Pain Res Manag [Internet]. 2015 Sep;20(5):255–60. Available from: http://ezproxy.bangor.ac.uk/login?url=http://search.ebscohost.com/login.aspx?direct=true&db=rzh&AN=110236791&site=ehost-live

28. Kaphle S, Hancock H, Newman LA. Childbirth traditions and cultural perceptions of safety in Nepal: critical spaces to ensure the survival of mothers and newborns in remote mountain villages. Midwifery. 2013 Oct 1;29(10):1173–81.

29. Keynan HA. Male roles and the making of the Somali tragedy. Reflections on gender, masculinity and violence in Somali society. InProceedings of EASS/SSIA International Congress of Somali Studies. Variations on the theme of Somaliness. 2001. Centre for Continuing Education, Åbo Akademi University.

30. Kumar KH, Elavarasi P. Definition of pain and classification of pain disorders. Journal of Advanced Clinical and Research Insights. 2016 May 1;3(3):87–90.

31. Lasch KE. Culture, pain, and culturally sensitive pain care. Pain management nursing. 2000 Sep 1;1(3):16–22.

32. Lee JS, Akhtar S. Effects of the workplace social context and job content on nurse burnout. Human Resource Management. 2011 Mar;50(2):227–45.

33. Lohman D, Schleifer R, Amon JJ. Access to pain treatment as a human right. BMC medicine. 2010 Dec;8(1):1–9.

34. Lourens A, Parker R, Hodkinson P. Prehospital acute traumatic pain assessment and management practices in the Western Cape, South Africa: a retrospective review. Int J Emerg Med [Internet]. 2020 May 5;13(1):1–10. Available from: http://ezproxy.bangor.ac.uk/login?url=http://search.ebscohost.com/login.aspx?direct=true&db=rzh&AN=143056750&site=ehost-live

35. Low D, Merkel EC, Menon M, Loggers E, Ddungu H, Leng M, Namukwaya E, Casper C. End-of-life palliative care practices and referrals in Uganda. Journal of Palliative Medicine. 2018 Mar 1;21(3):328–34.

36. Miftah R, Tilahun W, Fantahun A, Adulkadir S, Gebrekirstos K. Knowledge and factors associated with pain management for hospitalized children among nurses working in public hospitals in Mekelle City, North Ethiopia: cross sectional study. BMC Res Notes. 2017;10(1):2–7.

37. Moher D, Shamseer L, Clarke M, Ghersi D, Liberati A, Petticrew M, Shekelle P, Stewart LA. Preferred reporting items for systematic review and meta-analysis protocols (PRISMA-P) 2015 statement. Systematic reviews. 2015 Dec;4(1):1–9.

38. Nabukenya MT, Kintu A, Wabule A, Muyingo MT, Kwizera A. Knowledge, attitudes and use of labour analgesia among women at a low-income country antenatal clinic. BMC Anesthesiol [Internet]. 2015 Jul;15(1):1–6. Available from: http://ezproxy.bangor.ac.uk/login?url=http://search.ebscohost.com/login.aspx?direct=true&db=rzh&AN=108665556&site=ehost-live

39. Nchako E, Bussell S, Nesbeth C, Odoh C. Barriers to the availability and accessibility of controlled medicines for chronic pain in Africa. International health. 2018 Mar 1;10(2):71–7.

40. Noah D, Fidas G. The global infectious disease threat and its implications for the United States. National Intelligence Council Washington DC; 2000 Jan 1.

41. Nortjé N, Albertyn R. The cultural language of pain: A South African study. South African Fam Pract [Internet]. 2015;57(1):24–7. Available from: http://dx.doi.org/10.1080/20786190.2014.977034

42. Nortjé N, Albertyn R. The cultural language of pain: A South African study. South African Fam Pract [Internet]. 2015;57(1):24–7. Available from: http://dx.doi.org/10.1080/20786190.2014.977034

43. Olaogun A, Ayandiran O, Olalumade O, Obiajunwa P, Adeyemo F. Knowledge and management of infants’ pain by mothers in Ile Ife, Nigeria. Int J Nurs Pract [Internet]. 2008 Aug;14(4):273–8. Available from: http://ezproxy.bangor.ac.uk/login?url=http://search.ebscohost.com/login.aspx?direct=true&db=rzh&AN=105559287&site=ehost-live

44. Onsongo LN. Barriers to Cancer Pain Management Among Nurses in Kenya: A Focused Ethnography. Pain Manag Nurs [Internet]. 2020 Jun;21(3):283–9. Available from: http://ezproxy.bangor.ac.uk/login?url=http://search.ebscohost.com/login.aspx?direct=true&db=rzh&AN=143365219&site=ehost-live

45. Peacock S, Patel S. Cultural influences on pain. Reviews in pain. 2008 Mar;1(2):6–9.

46. Ratshikana-Moloko M, Ayeni O, Tsitsi JM, Wong ML, Jacobson JS, Neugut AI, et al. Spiritual Care, Pain Reduction, and Preferred Place of Death Among Advanced Cancer Patients in Soweto, South Africa. J Pain Symptom Manag [Internet]. 2020 Jul;60(1):37–47. Available from: http://ezproxy.bangor.ac.uk/login?url=http://search.ebscohost.com/login.aspx?direct=true&db=rzh&AN=143860147&site=ehost-live

47. Sonqishe M, Levy L. Pain control in a patient with myeloma in Zimbabwe… international feature. Cancer Nurs [Internet]. 1990 Jun;13(3):198–200. Available from: http://ezproxy.bangor.ac.uk/login?url=http://search.ebscohost.com/login.aspx?direct=true&db=rzh&AN=107521488&site=ehost-live

48. Suleiman ZA, Wahab KW, Kolawole IK. Opioid prescribing habits of physicians in Kwara State, Nigeria. Ghana Med J. 2016;50(2):63–7.

49. Toh SG, Ang E, Devi MK. Systematic review on the relationship between the nursing shortage and job satisfaction, stress and burnout levels among nurses in oncology/haematology settings. International Journal of Evidence-Based Healthcare. 2012 Jun;10(2):126–41.

50. Ufashingabire CM, Nsereko E, Njunwa KJ, Brysiewicz P. Knowledge and attitudes of nurses regarding pain in the intensive care unit patients in Rwanda. Rwanda Journal. 2016 Nov 1;3(1):21–6.

51. Woolley RJ, Velink A, Phillips RO, Thompson WA, Mohammed Abass K, Van Der Werf TS, et al. Experiences of pain and expectations for its treatment among former buruli ulcer patients. Am J Trop Med Hyg. 2016;95(5):1011–5.

52. Young JR, Sih C, Hogg MM, Anderson-Montoya BL, Fasano HT. Qualitative Assessment of Face Validity and Cross-Cultural Acceptability of the Faces Pain Scale: “Revised” in Cameroon. Qual Health Res [Internet]. 2018 Apr;28(5):832–43. Available from: http://ezproxy.bangor.ac.uk/login?url=http://search.ebscohost.com/login.aspx?direct=true&db=rzh&AN=128684332&site=ehost-live

53. Young JR, Sih C, Hogg MM, Anderson-Montoya BL, Fasano HT. Qualitative Assessment of Face Validity and Cross-Cultural Acceptability of the Faces Pain Scale: “Revised” in Cameroon. Qual Health Res [Internet]. 2018 Apr;28(5):832–43. Available from: http://ezproxy.bangor.ac.uk/login?url=http://search.ebscohost.com/login.aspx?direct=true&db=rzh&AN=128684332&site=ehost-live

